# Test re-test reliability and minimal detectable change of pulse wave velocity and flow mediated dilation in paretic and non-paretic limbs of older adults post-stroke

**DOI:** 10.64898/2026.09.18.26363408

**Authors:** Juliano A Abreu, Natalie D’Isabella, Derek W Stouth, Maureen MacDonald, Ada Tang, Kevin Moncion

## Abstract

Arterial stiffness and endothelial dysfunction are non-traditional risk markers after stroke, yet reliability and minimal detectable change (MDC) of pulse wave velocity (PWV) and flow-mediated dilation (FMD) have not been established. We examined test-retest reliability and MDC of carotid-femoral (cfPWV), carotid-radial (crPWV), and femoral-dorsalis pedis PWV (ffPWV), and absolute and relative brachial artery FMD in people >12 months post-stroke. Seventeen individuals (n=4(24%) females; 63.9±8.6 years; 4.5±2.8 years post-stroke) completed two bilateral assessments of PWV via applanation tonometry and FMD via duplex ultrasound within one week. Intraclass correlation coefficients (ICC_2,1_) were calculated via two-way random-effects models, and MDC derived at 68%, 90%, and 95% confidence levels. We found good-to-excellent reliability for cfPWV in paretic (ICC_2,1_=0.85; 95%CI:0.59,0.95) and non-paretic (ICC_2,1_=0.94; 95%CI:0.82,0.98) limbs, with MDC_95%_ values of 2.23 and 1.68 m/s, respectively. crPWV and ffPWV exhibited poor reliability bilaterally (ICC_2,1_=0.12–0.33). Relative FMD showed-moderate reliability for paretic (ICC_2,1_=0.50; 95%CI:-0.20,0.85) and non-paretic (ICC=0.59; 95%CI:0.04,0.84) limbs, with MDC_95%_ values of 6.88% and 7.65%, respectively. Reliability for absolute FMD was poor (ICC_2,1_=0.11–0.44). These findings support cfPWV as a reliable vascular assessments in individuals >12 months post-stroke, and provide stroke-specific MDC values to guide interpretability in future trials.

## Introduction

Stroke is among the leading causes of death and long-term disability worldwide, with survivors facing substantially elevated risk of recurrent stroke and cardiovascular-related events.^1,2^ Although the combined population attributable fraction of stroke risk due to traditional cardiovascular risk factors (i.e., hypertension, hyperlipidemia, hyperglycemia) is roughly 85%,^3^ non-traditional risk markers including arterial stiffness and endothelial dysfunction may provide additional prognostic information regarding cardiovascular disease development and progression.^4,5^ Carotid-femoral pulse wave velocity (cfPWV), considered the gold-standard measure of central arterial stiffness, reflects the speed at which pulse pressure waves propagate in opposite directions to the carotid and femoral arterial sites.^6,7^ Carotid-radial and femoral-dorsalis pedis arterial sites have also been used to assess upper and lower body peripheral arterial stiffness, respectively. Together the assessment of different arterial sites may provide a more informed measurement of varying structural and histological changes in stiffness.^8^ Endothelial function is commonly assessed via flow-mediated dilation (FMD) and provides information regarding the functional integrity of the vascular endothelium.^5^ Both arterial stiffness and endothelial dysfunction have emerged as factors associated with stroke incidence^9–12^ and stroke severity,^11^ and appear to be impaired across all phases of stroke recovery.^13^

Changes in structural and functional vascular adaptations occur in the paretic limb after stroke which may influence blood flow during exercise and rehabilitation. Reduced arterial diameter, blood flow and oxygen consumption, and impairments in microvascular function have been reported in the paretic limb relative to the non-paretic limb in the acute, subacute and chronic phases of stroke recovery.^14–18^ These inter-limb differences may be due in part to reduced metabolic demand (e.g., oxygen consumption),^17^ neural coupling,^19^ and skeletal muscle fiber characteristics in the paretic limb.^20^ Furthermore, hemispheric lateralization of stroke may differentially influence endothelial function and arterial compliance, with individuals with right-hemisphere stroke demonstrating reduced vascular function compared to those with left-sided involvement.^21^ Whether these inter-limb vascular differences influence measurement properties between paretic and non-paretic limbs has not yet been explored in people post-stroke.

Establishing measurement properties for vascular outcomes after stroke can inform the selection of outcome measures and the interpretation of findings from intervention studies. Relative reliability quantifies the extent to which individuals maintain their rank order across repeated measurements (e.g., test-retest, intra-rater reliability using intra-class correlation coefficients [ICC]),^22^ absolute reliability quantifies the extent to which repeated measures vary within individuals (e.g., standard error of measurement, [SEM]),^22^ and minimal detectable change (MDC) represents the smallest absolute change that exceeds measurement error within a specified level of confidence (e.g., MDC_95%_), allowing researchers to distinguish true physiological change that is beyond measurement error.^23^ Test-retest reliability of PWV has been established in the general population^24^ and in individuals with spinal cord injury.^25^ Among adults aged 45-64 years old, variable ICCs and MDCs have been reported that differ across arterial segments, with cfPWV ICC = 0.70 [95% CI 0.59, 0.81] and MDC ≈ 4.1 m/s; versus femoral-dorsalis pedis PWV ICC = 0.69 [95% CI 0.59, 0.79] and MDC ≈ 3.0 m/s.^24^ Despite the availability of standardized protocols for the assessment of FMD,^5^ measured values appear to be variable in non-stroke (i.e., coefficients of variation ranging between 15.8 to 17.5%)^26^ and stroke populations.^13^ Given the common presentation of unilateral impairment after stroke, whether there are differences in absolute and relative reliability of vascular measures between paretic and non-paretic limbs have also not yet been evaluated.

Therefore, the purpose of this study was to examine the relative test-retest reliability, absolute reliability, and the MDC of PWV at commonly measured arterial segments (cfPWV, carotid-radial PWV (crPWV), femoral-dorsalis pedis PWV (ffPWV)), and relative and absolute brachial artery FMD in paretic and non-paretic limbs in individuals with stroke.

## Methods

### Study Design

This was an observational, cross-sectional study with repeated measures within 1 week. Informed written consent was obtained from all study participants. This study was approved by the Hamilton Integrated Research Ethics Board (#13-348).

### Participants

Participants were recruited from the community within the Ontario Central South Stroke Network (Hamilton, Ontario), and were contacted via phone or email if they had previously consented to be contacted for future studies. Participants were eligible for the study if they were 50-80 years old, ≥ 12 months post-stroke, and able to walk at least 10 meters independently with or without an assistive device. Participants were excluded if they had stroke of non-cardiogenic origin, uncontrolled arrhythmias (Class C or D American Heart Association Risk Criteria), hypertension (resting blood pressure > 160/100 mmHg), significant musculoskeletal problems (rheumatoid arthritis), significant neurological conditions (e.g., Parkinson’s), co-morbidities (e.g., unclipped aneurysms, uncontrolled seizures etc.), cognitive or communication impairment, or behavioral issues affecting ability to understand instructions. Participant enrollment occurred between 2013 and 2017.^27^

### Assessments

Demographic information including age, biological sex, medical history, and anthropometric measurements (weight, height, body mass index) were collected for all participants. Functional capacity was assessed using the 6-minute walk test. Information regarding timing post-stroke, stroke type, and stroke severity as assessed by the National Institutes of Health Stroke Severity Scale (NIHSS)^28^ and Chedoke-McMaster Stroke Assessment (CMSA)^29^ were collected. Global cognitive function was assessed using the Montreal Cognitive Assessment (MoCA).^30^

### Vascular Experimental Procedures

All vascular assessments were conducted during two visits at the same time of day scheduled within a one-week period in a temperature-controlled laboratory (23°C). PWV and FMD assessments were conducted by trained assessors following standard operating procedures. Prior to each visit, participants fasted from food or drink for four hours, abstained from caffeine for 8 hours and smoking for 12 hours, and avoided structured physical activity for 24 hours. Participants were instructed to take medications as prescribed.

Upon arrival at the laboratory, participants were instrumented with a 3-lead electrocardiogram (Dual Bio Amp FE232, ADInstruments, Bella Vista, New South Wales, Australia) and a non-invasive finger blood pressure monitoring system (Finometer MIDI, Finapres Medical Systems, Amsterdam, Netherlands) to measure continuous heart rate, systolic and diastolic blood pressure and mean arterial pressure over a 10-minute supine resting period. Blood pressure was also assessed at the level of the brachial artery (Dinamap V100, General Electric Healthcare Inc, Chicago, IL, United States). Two discrete readings were measured and averaged. If values differed by > 5 mmHg, 2 additional readings were taken and the average across 4 readings was used.

Following the rest period, arterial stiffness was assessed non-invasively in supine using applanation tonometry (SPT-301, Millar Instruments Inc., Houston, TX, United States) connected to a pressure control unit (Millar Instruments Inc). Two hand-held microtip pressure sensitive tonometers (model SPT-301. Millar Instruments Inc.) were connected to a pressure control unit (Millar Instruments Inc.) and all pressure signals were collected sequentially at 200 Hz using an analogue-to-digital data acquisition system (PowerLab, ADInstruments, Colorado Springs, CO, United States). Pressure waveforms were band-pass filtered (5-30 Hz) and the foot of each waveform was identified as the minimum value of the filtered signal (LabChart7 Pro, ADInstruments, Colorado Springs, CO, United States).^31^ Pulse waveforms were calculated with simultaneous waveform detection at the distal and proximal locations across the carotid-femoral (cfPWV), carotid-radial (crPWV), and femoral-dorsalis pedis (ffPWV) arterial segments. Distance measurements were obtained after each pressure waveform data collection phase using a straight and taut tape measure over the surface of the body. cfPWV distance was calculated as the distance from the sternal notch to the umbilicus to the femoral artery measurement site minus the distance from the sternal notch to the carotid artery measure site. crPWV distance was calculated as the distance from the sternal notch to the radial artery measurement site, with the arm abducted 30 degrees from the body, minus the distance from the sternal notch to the carotid artery site. ffPWV distance was calculated as the distance from the femoral artery measurement site to the dorsalis pedis artery measurement site.

PWV was calculated as the distance (meters)/Δt (seconds), where Δt represented the time delay between the foot of the two waveforms (e.g., carotid and femoral). PWV values were averaged across 30 consecutive cardiac cycles. All PWV measurements were collected twice on both paretic and non-paretic sides.

Brachial artery FMD was measured via Duplex mode ultrasound (Vivid q; GE Medical Systems, Horten, Norway) with a 12 MHz linear array probe at 7.7 fps on both the paretic and non-paretic sides following current guidelines.^4^ Each FMD test included brachial artery ultrasound imaging over 3 phases: 1) a 30-second resting phase to capture baseline vessel diameter, 2) a 5-minute ischemic occlusion phase, which involved the rapid inflation of a sphygmomanometer cuff to 200 mmHg (E20 Rapid Cuff Inflator and AG101 Air Source; Hokanson, Bellevue, WA, United States) applied distal to the cubital fossa, and 3) a reactive hyperemia phase, where the cuff was rapidly deflated and the brachial artery imaged for 3 more minutes to capture peak arterial diameter. All ultrasound images were saved and analyzed using semiautomated edge-tracking software (Arterial Measurement System, Gothenburg, Sweden). Baseline vessel diameter was calculated as the 30-second resting phase mean. During post-deflation reactive hyperemia, arterial diameter was smoothed using five-cardiac-cycle moving average, and peak diameter was defined as the maximum of these moving averages. Absolute FMD was calculated as the difference between the peak and baseline diameters [FMD (mm) = peak diameter-baseline diameter], while relative FMD was calculated using allometric scaling, defined as the ratio of peak artery diameter relative to baseline diameter raised to the population-specific allometric scaling exponent [FMD (%) = (peak diameter / baseline diameter ^ scaling unit) * 100%]. Relative FMD calculated as the percent change in artery diameter relative to baseline diameter [FMD (%) = (peak diameter – baseline diameter) / baseline diameter) * 100%) is reported in the supplementary material (Appendix A).

### Statistical Analysis

Participant demographic statistics were summarized as means (standard deviation) or medians (interquartile ranges [IQR]) for normally and non-normally distributed data, respectively, and frequencies (n, %) for categorical data. The ICC with a two-way random-effects model and single-rater absolute agreement (ICC_2,1_) was used to evaluate the relative measurement reliability of each arterial segment and the paretic and non-paretic limbs. ICC parameters were reported and interpreted according to guidelines for selecting and reporting ICCs in reliability research.^32^ ICC values greater than 0.90 indicate excellent reliability, between 0.90 and 0.75 indicate good reliability, between 0.5 and 0.75 indicate moderate reliability, and lower than 0.5 indicate poor reliability.^32^ The SEM was used for absolute reliability, and the MDC was used to estimate true statistical change at the 68% (MDC68), 90% (MDC90), and 95% (MDC95) confidence levels. Bland-Altman plots were used to display the limits of agreement for each measurement between the paretic and non-paretic sides.^32,33^ All reliability and agreement analyses were restricted to participants who completed both testing sessions; individuals with missing data were not included in site-specific analyses. All statistical analyses were conducted using Stata 19 (StataCorp, College Station, Texas, United States) and figures were created with R (v4.4.2; R Core Team, Vienna, Austria).

## Results

In total, 17 (n=4 females) participants were included in the current analysis. Participant demographics are reported in Table 1. Participants were 63.9 (8.6) years old, were 4.5 (2.8) years post-stroke, had a body mass index of 31.2 (5.5) kg/m^2^. They had a 6-minute walk test time of 398.35 (108.46) meters, NIHSS score of 2 [2] indicating mild severity stroke, MoCA score of 22 [4] indicating mild cognitive impairment, CMSA Arm score of 5 [2] indicating reduced spasticity that is evident with rapid movements and at extremes ranges. Resting heart rate was 66 (13) beats per minute, resting systolic blood pressure 136 (17) mmHg, and resting diastolic blood pressure 80 (10) mmHg.

**Table 1.** Participant demographics (N=17)

| Variable | Value |
| --- | --- |
| Age, years | 63.9 (8.6) |
| Females, n (%) | 4 (23.5%) |
| Weight, kg | 89.9 (18.5) |
| Height, cm | 169.6 (8.6) |
| Body mass index, kg/m<sup>2</sup> | 31.2 (5.5) |
| 6-minute walk test, m | 398.35 (108.46) |
| National Institute of Health Stroke Scale (median, [IQR]) | 2 [2] |
| Montreal Cognitive Assessment (median, [IQR]) | 22 [4] |
| CMSA Arm (median, [IQR]) | 5 [2] |
| Time post-stroke, years | 4.5 (2.8) |
| Comorbidities, n (median, [IQR]) | 3 [1] |
| Hypertension, n (%) | 5 (31.3%) |
| Antihypertensive medication, n (%) | 5 (33.3%) |
| Resting heart rate, beats/minute | 66 (13) |
| Resting systolic blood pressure, mmHg | 136 (17) |
| Resting diastolic blood pressure, mmHg | 80 (10) |

Values are mean (standard deviation) or medians (interquartile ranges [IQR]) for normally and non-normally distributed data, respectively, and frequencies (n, %) for count data.

### Relative and Absolute Reliability of Pulse Wave Velocity

Test scores and reliability values for PWV at each arterial segment and by paretic and non-paretic limbs are presented in Table 2. Bland-Altman plots and the level of agreement for each measurement between the paretic and non-paretic sides is presented in Figure 1.

**Figure 1.**
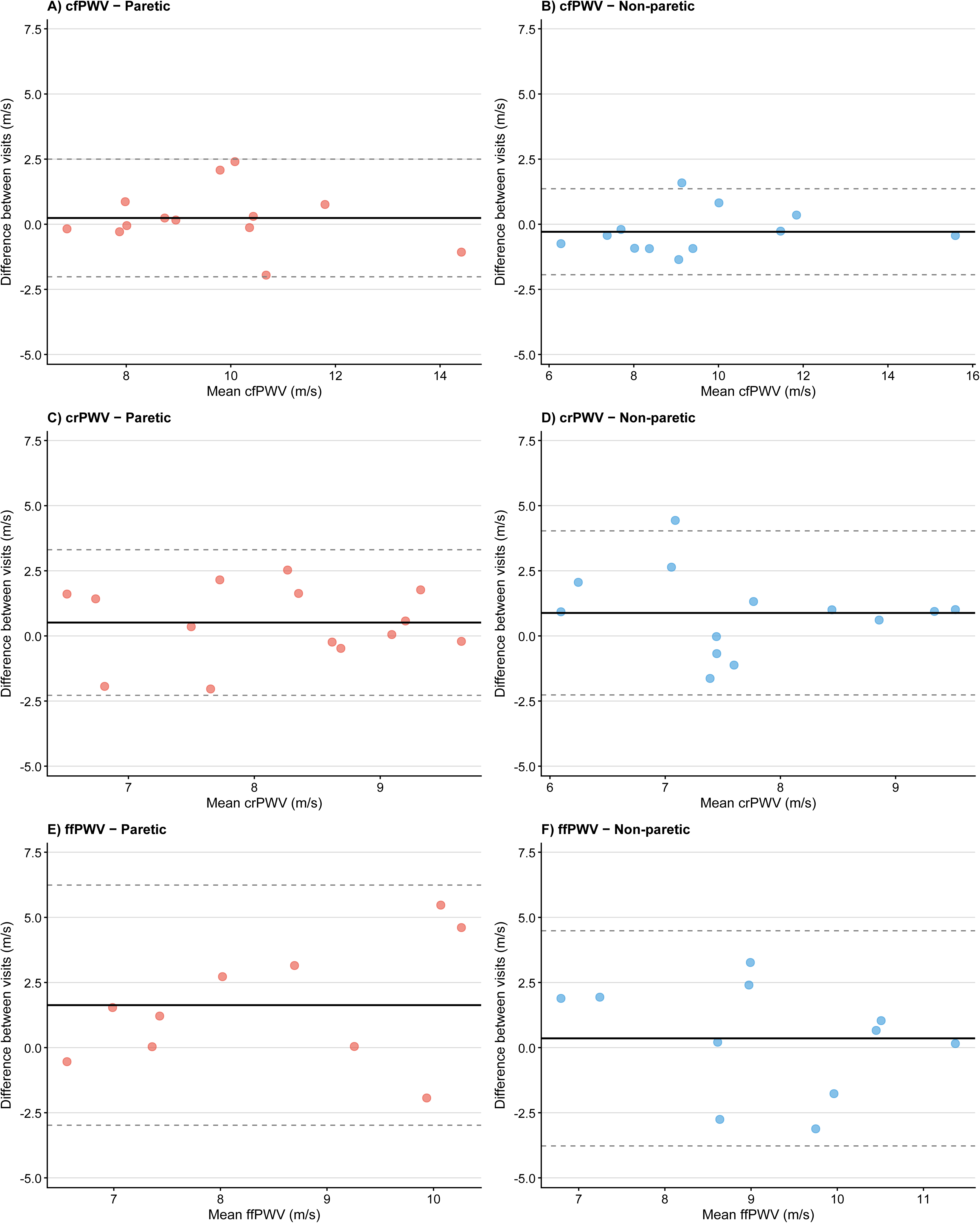
Bland-Altman plots demonstrating limits of agreement and absolute test-retest reliability by paretic (red) and non-paretic (blue) sides for carotid-femoral (N=13,12), carotid-radial (N=14,13), and femoral-dorsalis pedis (N=10,11) PWVarterial segments. The black solid line represents the mean difference between visits and the dashed lines represents the limits of agreement.

**Table 2.** Relative and Absolute Reliability of PWV and FMD by paretic and non-paretic limbs.

| Measures | N | Test 1 (SD) | Test 2 (SD) | SD<sub>Pooled</sub> | ICC<sub>2,1</sub> (95% CI) | SEM | MDC<sub>68%</sub> | MDC<sub>90%</sub> | MDC<sub>95%</sub> |
| --- | --- | --- | --- | --- | --- | --- | --- | --- | --- |
| **cfPWV** |  |  |  |  |  |  |  |  |  |
| Paretic | 13 | 9.81 (1.97) | 9.57 (2.16) | 2.07 | 0.85 (0.59, 0.95) | 0.80 | 1.14 | 1.87 | 2.23 |
| Non-Paretic | 12 | 9.38 (2.61) | 9.66 (2.45) | 2.53 | 0.94 (0.82, 0.98) | 0.61 | 0.86 | 1.41 | 1.68 |
| **crPWV** |  |  |  |  |  |  |  |  |  |
| Paretic | 14 | 8.41 (1.26) | 7.90 (1.23) | 1.24 | 0.33 (-0.18, 0.71) | 1.02 | 1.44 | 2.37 | 2.82 |
| Non-Paretic | 13 | 8.16 (1.22) | 7.27 (1.43) | 1.33 | 0.24 (-0.22, 0.66) | 1.17 | 1.65 | 2.71 | 3.23 |
| **ffPWV** |  |  |  |  |  |  |  |  |  |
| Paretic | 10 | 9.27 (2.13) | 7.64 (1.42) | 1.81 | 0.12 (-0.30, 0.61) | 1.70 | 2.40 | 3.95 | 4.71 |
| Non-Paretic | 11 | 9.39 (1.47) | 9.03 (1.98) | 1.74 | 0.28 (-0.39, 0.74) | 1.48 | 2.09 | 3.44 | 4.09 |
| **Absolute FMD** |  |  |  |  |  |  |  |  |  |
| Paretic | 10 | 0.19 (0.07) | 0.19 (0.14) | 0.11 | 0.11 (-0.63, 0.69) | 0.10 | 0.15 | 0.24 | 0.29 |
| Non-Paretic | 10 | 0.12 (0.18) | 0.20 (0.14) | 0.16 | 0.44 (-0.13, 0.81) | 0.12 | 0.17 | 0.28 | 0.34 |
| **Relative FMD** |  |  |  |  |  |  |  |  |  |
| Paretic | 10 | 6.43 (4.04) | 4.43 (6.73) | 5.55 | 0.50 (-0.20, 0.85) | 2.48 | 3.51 | 5.77 | 6.88 |
| Non-Paretic | 10 | 4.76 (6.54) | 7.02 (0.578) | 6.17 | 0.59 (0.04, 0.84) | 2.76 | 3.90 | 6.42 | 7.65 |

Abbreviations. cfPWV: carotid-femoral pulse wave velocity; crPWV: carotid-radial pulse wave velocity; ffPWV: femoral-dorsalis pedis pulse wave velocity; FMD: flow mediated dilation; ICC<sub>2,1</sub>: intra-class correlation coefficient; MDC: minimal detectable change; SEM: standard error of measurement.

PWV measurement distances (m) were consistent between repeated measurements on the paretic and non-paretic sides (paretic test 1, paretic test 2, non-paretic test 1, non-paretic test 2): cfPWV: 0.52 (0.06), 0.53 (0.06), 0.51 (0.07), 0.53 (0.05); crPWV: 0.56 (0.04), 0.57 (0.05), 0.58 (0.04), 0.56 (0.04); ffPWV: 0.85 (0.08), 0.82 (0.05), 0.83 (0.06), 0.81 (0.05).

The cfPWV values demonstrated good to excellent relative reliability for both paretic (ICC = 0.85; 95% CI 0.59, 0.95, N=13) and non-paretic (ICC = 0.94; 95% CI 0.82, 0.98, N=12) limbs. The SEM was 0.80 m/s for paretic and 0.61 m/s for non-paretic limbs with corresponding to MDC_95%_ values of 2.23 m/s and 1.68 m/s, respectively.

The crPWV values demonstrated poor relative reliability for both paretic (ICC = 0.33; 95% CI -0.18, 0.71, N=14) and non-paretic (ICC = 0.24; 95% CI -0.22, 0.66, N=13) limbs. The SEM was 1.02 m/s for paretic and 1.17 m/s and non-paretic limbs, with MDC_95%_ values of 2.82 m/s and 3.23 m/s, respectively.

The ffPWV demonstrated poor relative reliability for both paretic (ICC = 0.12; 95% CI -0.30, 0.61, N=10) and non-paretic (ICC = 0.28; 95% CI -0.39, 0.74, N=11) limbs. The SEM was 1.70 m/s for paretic and 1.48 m/s for non-paretic limbs, with MDC_95%_ values of 4.71 m/s and 4.09 m/s, respectively.

### Absolute and Relative Reliability of Flow-Mediated Dilation

Test scores and reliability values for FMD by paretic and non-paretic limbs are presented in Table 2. Baseline, 4-minute occlusion, and reactive hyperemia vessel diameters are presented in Table 3. Bland-Altman plots demonstrating the limits of agreement and absolute test-retest reliability for absolute and relative FMD and between the paretic and non-paretic sides are presented in Figure 2.

**Figure 2.**
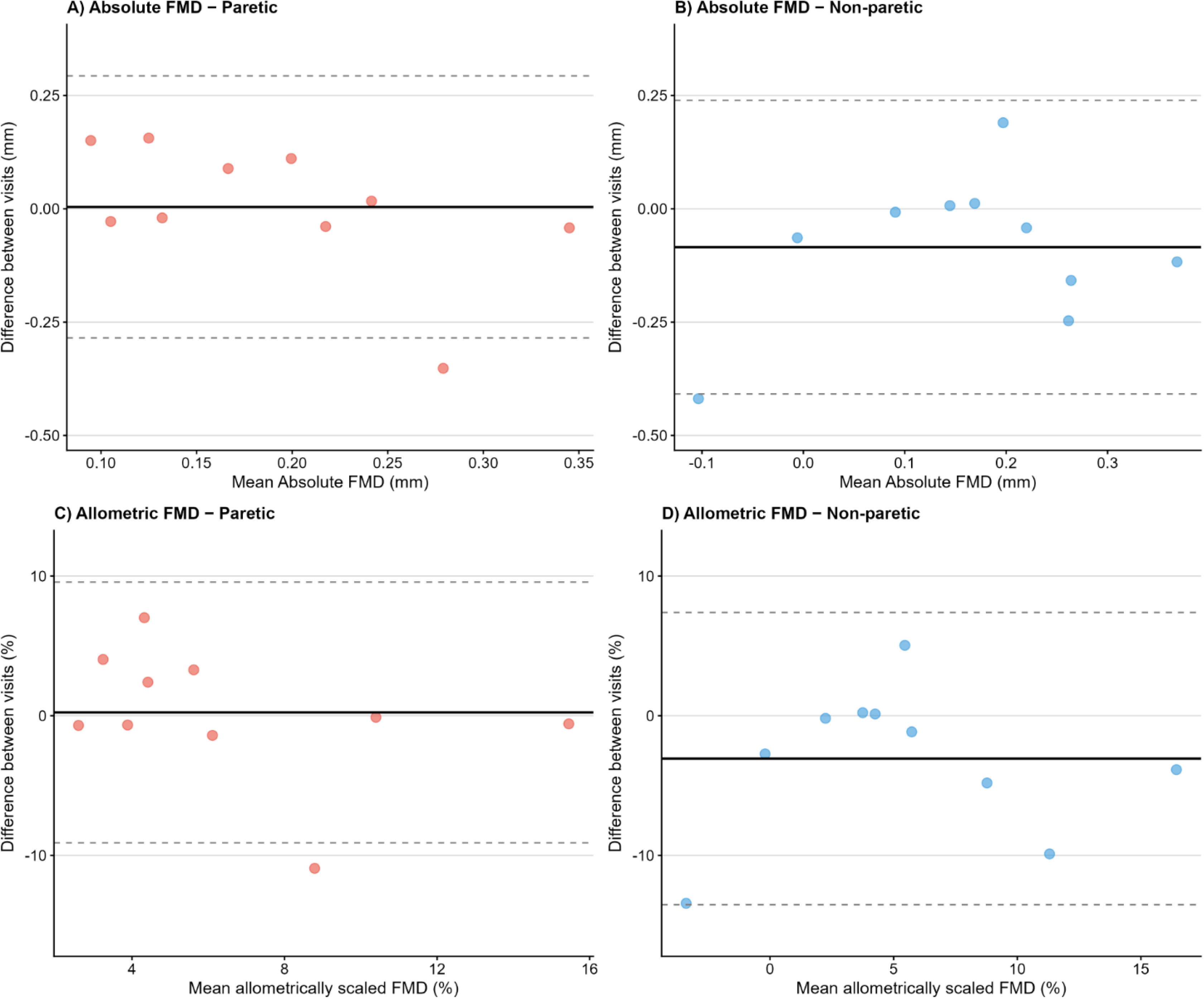
Bland-Altman plots demonstrating limits of agreement and absolute test-retest reliability for absolute FMD and relative FMD by paretic and non-paretic sides (N=10). The black solid line represents the mean difference between visits and the dashed lines represents the limits of agreement.

**Table 3.** Flow-mediated dilation measurements across tests.

| Measures | N | Test 1 (SD) | Test 2 (SD) | Mean |
| --- | --- | --- | --- | --- |
| Baseline Diameter | 10 | 3.75 (1.01) | 3.79 (0.96) | 3.77 (0.98) |
| 4-Minute Occlusion Diameter | 10 | 3.74 (1.02) | 3.83 (0.96) | 3.79 (0.98) |
| Reactive Hyperemia Diameter | 10 | 3.92 (1.00) | 3.97 (0.94) | 3.95 (0.96) |

Absolute FMD (mm) demonstrated poor reliability for both paretic (ICC = 0.11; 95% CI −0.63, 0.69, N=10) and non-paretic (ICC = 0.44; 95% CI −0.13, 0.81, N=10) limbs. The SEM was 0.10 mm for paretic and 0.12 mm for non-paretic limbs, with MDC_95%_ values of 0.29 mm and 0.34 mm, respectively.

Relative FMD (%) demonstrated moderate reliability for the paretic (ICC = 0.50; 95% CI -0.20, 0.85, N=10) and non-paretic limbs (ICC = 0.59; 95% CI 0.04, 0.84, N=10). The SEM was 2.48% for paretic and 2.74% non-paretic limbs, with MDC_95%_ values of 6.88% and 7.65%, respectively.

## Discussion

The current study examined the relative and absolute reliability of PWV and FMD in stroke. Our findings suggest that cfPWV may be a reliable measure for PWV in individuals >12 months post-stroke, with paretic (ICC = 0.85) and non-paretic limbs (ICC = 0.94) exhibiting good and excellent test-retest reliability, respectively. Peripheral segments of crPWV and ffPWV however exhibited poor test-retest reliability in both paretic and non-paretic sides following stroke. Our results also demonstrated moderate reliability for relative FMD in paretic (ICC = 0.50) and non-paretic limbs (ICC = 0.59), respectively, and poor reliability for absolute FMD in both conditions. As such, there were no key differences in reliability metrics for relative FMD between paretic and non-paretic limbs. Taken together, our findings suggest that inter-limb vascular adaptations >12 months post-stroke^14–18^ may not appreciably influence measurement properties for these commonly referenced vascular outcomes. Thus, in accordance with standard methodological practices for PWV and FMD^5,7^ future studies after stroke may consider prioritizing cfPWV and relative FMD measures collected on the right side.

We provide stroke-specific evidence that supports cfPWV as the gold-standard assessment of central arterial stiffness,^7^ given its superior reliability to peripheral PWV measurements in this population. These findings are consistent with reliability metrics reported in non-stroke older adults (ICC = 0.70)^24^ and individuals with spinal cord injury (ICC = 0.92 to 0.98).^25^ The poor reliability in peripheral PWV measures seen in the current study may be partly explained by the anatomical and physiological features of peripheral arteries and cardiovascular risk factors that are commonly present post-stroke. For instance, smaller peripheral arteries such as the radial and dorsalis pedis, are predominantly muscular in structure, have larger wall-to-lumen ratios, and tend to be more responsive to vasodilatory stimuli relative to larger vessels^34^. Following stroke, there are changes in muscle sympathetic nerve activity^35^ and increased levels of muscle spasticity and tone in the periphery.^36^ These factors are further compounded by the cardiovascular risk profile of individuals post-stroke who are more likely to have hypertension, type-2 diabetes and obesity, all of which are risk factors for endothelial dysfunction post-stroke^18,37^. The convergence of these factors may lead to greater variability and thus measurement error in crPWV and ffPWV as indicators of peripheral arterial stiffness. Our findings therefore support cfPWV, but not peripheral PWV, as a potentially reliable measurement outcome of central arterial stiffness in stroke.

Our findings also provide evidence of moderate reliability in relative FMD for the paretic (ICC = 0.50; 95% CI: -0.20, 0.85) and non-paretic (ICC = 0.59; 95% CI: 0.04, 0.84) limbs, whereas absolute FMD was poor in both limbs (ICC = 0.11–0.44). Although baseline and peak diameter measurements appeared consistent across tests, it is possible that variability in baseline measurements were negated in relative but not absolute measures, contributing to increased error and decreased reliability for absolute FMD assessments. We note however the wide confidence intervals around the point estimates for relative FMD, reflecting imprecision that may be due to the small sample size and warrants careful interpretation but appears to be consistent with other research. The relative FMD values in our sample (4-7%) are consistent with the pooled estimate from a recent meta-analysis by Bartsch et al. (2024), who reported a mean FMD of 3.9% (95% CI: 2.5–5.3%) albeit also with extreme heterogeneity (I^2^ = 99%, range -4% to 13%) across 28 studies post-stroke^13^. Of interest, a meta-regression controlling for age and time-post stroke found little to no influence on the reported between-study heterogeneity, ^13^suggesting that methodological considerations rather than participant characteristics likely contribute to this source of heterogeneity. Indeed, adhering to gold-standard guidelines, including experienced sonographers, static probe holders, and automated edge-detection software is needed to further improve the precision of FMD estimates in future stroke research.^5^

We also provide absolute measures of reliability to guide the interpretation of true statistical change in vascular outcomes after stroke. The MDC_95%_ for cfPWV was 2.23 m/s and 1.68 m/s, while the MDC_95%_ relative FMD was 6.88% and 7.65% on the paretic side and on the non-paretic side, respectively. MDCs in cfPWV were lower when compared to other non-stroke populations at risk of atherosclerosis (MDC_95%_ ≈ 4.1 m/s). ^21^ These differences may be due to timing differences between measurements. In the ARIC study,^21^ repeated measurements were conducted 4-8 weeks apart, whereas measurements in the current study were repeated within 1 week to limit the likelihood that observed differences reflect true physiological change rather than measurement error. Conversely, MDCs in relative FMD in the current sample were higher than established MDCs in post-menopausal women (relative FMD = 2.92%)^38^ and estimated MDCs (1.5-2%) in older adults.^39^ It is possible that stroke specific changes in vascular function (i.e., muscle sympathetic nerve activity, muscle spasticity) and a relatively small sample size may have contributed to increased variability between subjects, thereby increasing MDCs. Taken together, timing of follow-up assessments, stroke severity, and sample size should all be considered when estimating minimal statistical differences in future research.

It is also important to note that the MDC does not correspond with the minimally clinically important differences (MCID). As such, our findings should not be used to determine whether observed changes are clinically meaningful, but rather interpreted in conjunction with statistical analyses to determine whether changes exceed expected measurement error. The most commonly referenced clinically meaningful threshold for PWV is derived from a meta-analysis of epidemiological data in non-stroke cohorts that demonstrate each 1 m/s increase in central PWV is associated with a ∼15% increase in cardiovascular event.^33^ Similarly, in the context of FMD, meta-analyses in non-stroke cohorts indicate 13% lower risk of cardiovascular events per percent point increase in brachial artery FMD.^5^ While these are undoubtedly clinically important, future research using anchor-based methods are needed to develop stroke-specific MCIDs for cfPWV and relative FMD to guide future clinical trial design and effect size calculations.

### Limitations

We acknowledge a few limitations in the current study. Although trained assessors followed standard operating procedures and gold-standard methods for the assessment of PWV and FMD,^5,7^ measurement error is unavoidable and, it is therefore possible that operator skill may have contributed to measurement variability between different assessment protocols. Additionally, our findings should be interpreted within the specific context of the current sample, which included individuals who were mostly male, more than 12 months post-stroke, and had mild stroke severity. Although participants were representative in terms of cardiovascular comorbidities, our results may not generalize to individuals in the acute phase of stroke or those with greater neurological severity. Further research is warranted examining reliability and MDCs across the continuum of recovery and severity. Finally, we advise caution when comparing assessments with more than one assessor as inter-rater reliability has not yet been examined in PWV and FMD within stroke populations.

## Conclusions

The current study is the first to our knowledge to examine test-retest reliability of PWV and FMD in individuals living with stroke. These preliminary findings suggest that cfPWV may be the most reliable measurements of arterial stiffness in individuals with chronic stroke and mild stroke severity. We provide stroke-specific reliability metrics to guide the interpretation of meaningful change in commonly beyond measurement error using MDC cutoffs for future stroke clinical trials; however, more work with greater sample sizes and more heterogeneity, especially with regard to gender, is needed for sufficient determination.

## Data Availability

All data produced in the present study are available upon reasonable request to the authors

