## Appendix A for "Test re-test reliability and minimal detectable change of pulse wave velocity and flow mediated dilation in paretic and non-paretic limbs of older adults post-stroke"

Appendix A. Relative and absolute reliability of relative FMD by paretic and non-paretic limbs.

| **Measures** | **N** | **Test 1 (SD)** | **Test 2 (SD)** | **$SD_{Pooled}$** | **$ICC_{2,1}$ (95% CI)** | **SEM** | **$MDC_{68\%}$** | **$MDC_{90\%}$** | **$MDC_{95\%}$** |
|---|---|---|---|---|---|---|---|---|---|
| Paretic | 10 | 5.88 (3.58) | 4.53 (4.34) | 3.98 | 0.78 (0.35, 0.94) | 1.86 | 2.63 | 4.32 | 5.15 |
| Non-Paretic | 10 | 4.21 (4.32) | 5.93 (5.63) | 5.02 | 0.70 (0.22, 0.92) | 2.74 | 3.87 | 6.36 | 7.58 |

Abbreviations. FMD: flow mediated dilation; $ICC_{2,1}$: intra-class correlation coefficient; MDC: minimal detectable change; SEM: standard error of measurement.
